# Novel Use of ctDNA to Identify Muscle-Invasive and Non-Organ Confined Upper Tract Urothelial Carcinoma

**DOI:** 10.1101/2023.03.28.23287866

**Authors:** Heather L Huelster, Billie Gould, Elizabeth A Schiftan, Lucia Camperlengo, Facundo Davaro, Kyle M Rose, Alex C Soupir, Shidong Jia, Tiantian Zheng, Wade J Sexton, Julio Pow-Sang, Philippe E Spiess, G. Daniel Grass, Liang Wang, Xuefeng Wang, Aram Vosoughi, Andrea Necchi, Joshua J Meeks, Bishoy M Faltas, Pan Du, Roger Li

**Author notes:** Corresponding Author: Roger Li, MD, Department of Genitourinary Oncology, H. Lee Moffitt Cancer Center & Research Institute 12901 USF Magnolia Drive, Tampa, FL 33612. Previously presented in poster format at ASCO Cancers Symposium 2022 on June 3, 2022 in Chicago, IL and podium format AUA National Meeting 2022 on May 16, 2022 in New Orleans, LA. Disclaimers and COI: Genomic sequencing supported by Predicine, Inc. Commercial funding sources did not play a role in the collection, interpretation of the data, writing of the report, or decision to submit the paper for publication.

## Abstract

**PURPOSE:** Optimal patient selection for neoadjuvant chemotherapy prior to surgical extirpation is limited by the inaccuracy of contemporary clinical staging methods in high-risk upper tract urothelial carcinoma (UTUC). We investigated whether the detection of plasma circulating tumor DNA (ctDNA) can predict muscle-invasive and non-organ confined (MI/NOC) UTUC.

**PATIENTS AND METHODS:** Plasma cell-free DNA was prospectively collected from chemotherapy-naïve, high-risk UTUC patients undergoing surgical extirpation and sequenced using a 152-gene panel and low-pass whole-genome sequencing. To test for concordance, whole exome sequencing was performed on matching tumor samples. The performance of ctDNA for predicting MI/NOC UTUC was summarized using area under a receiver-operating curve and the optimal variant count threshold determined using Younden’s J statistic. Kaplan-Meier methods estimated survival, and Mantel-Cox log-rank testing assessed the association between preoperative ctDNA positivity and clinical outcomes.

**RESULTS:** Of 30 patients prospectively enrolled, 14 were found to have MI/NOC UTUC. At least one ctDNA variant was detected from 21/30 (70%) patients with 52% concordance with matching tumor samples. Detection of at least two panel-based molecular alterations provided the optimal sensitivity and specificity to predict MI/NOC UTUC. Imposing this threshold in combination with a plasma copy number burden score >6.5 achieved a sensitivity of 79% and specificity of 94% in predicting MI/NOC UTUC. Furthermore, the presence of ctDNA was strongly prognostic for progression-free survival (1-yr PFS 69% vs. 100%, p<0.01) and overall survival (1-yr OS 56% vs. 100%, p<0.02).

**CONCLUSION:** The detection of plasma ctDNA prior to extirpative surgery was highly predictive of MI/NOC UTUC and strongly prognostic of PFS and OS. Preoperative ctDNA demonstrates promise as a biomarker for selecting patients to undergo neoadjuvant chemotherapy prior to nephroureterectomy.

## INTRODUCTION

Upper tract urothelial carcinoma (UTUC) is an aggressive disease with up to 70% incidence of high-grade histology and 60% muscle-invasive staging at the time of radical nephroureterectomy (RNU).^1^ Patients with muscle-invasive UTUC (≥pT2) have a poor prognosis, with 5-year cancer-specific mortality rates ranging between 21-59%.^2^ Fortunately, there is emerging evidence that cisplatin-based neoadjuvant chemotherapy (NAC) can be safely delivered to achieve pathologic down-staging and improved survival.^3,4^ However, a major challenge preventing optimal patient selection for NAC rests with the difficulty of accurate clinical staging due to UTUC’s cloistered anatomical location. Tumor biopsies by ureteroscopy under-stage UTUC up to 46% of the time.^5^ Efforts to improve clinical risk stratification using nomograms incorporating ureteroscopic findings, histologic features and cross-sectional imaging yielded only incremental gains.^6-10^ Moreover, clinical under-staging causes missed opportunities for systemic therapy, as those patients who develop renal insufficiency after surgery can no longer qualify for chemotherapy.^11^ Given the high stakes for accurate preoperative risk stratification, predictive biomarkers for invasive UTUC are critically needed.

The detection of circulating tumor DNA (ctDNA), that is, plasma cell-free DNA with tumor-specific alterations, is increasingly adopted for numerous clinical applications including cancer diagnosis, assessment of treatment response, and detection of residual disease and/or recurrence.^12,13^ ctDNA can be detected in up to 35% of patients with localized urothelial carcinoma of the bladder and 83% with metastatic urothelial cancer.^14-19^ Saliently, higher levels of ctDNA have been shown to correlate with disease burden and portend worse outcomes.^20^ In a proof-of-concept study, Birkenkamp-Demtroder et al^21^ demonstrated higher levels of ctDNA in patients with muscle invasive bladder cancer than those with recurrent non-muscle invasive disease. Based on these findings, we hypothesized that the detection of plasma ctDNA can be used to refine clinical staging in high-risk UTUC patients undergoing extirpative surgery. In this prospective observational study, we demonstrate the feasibility of preoperative ctDNA collection and correlate its accuracy in the prediction of muscle-invasive and non-organ confined UTUC (MI/NOC UTUC).

## PATIENTS AND METHODS

### Study Design, Patient Selection, and Clinical Sample Collection

Following IRB approval, 32 patients diagnosed with clinically resectable, high-risk UTUC and planning up-front RNU or ureterectomy at H. Lee Moffitt Cancer Center were prospectively enrolled from October 2020 to April 2022. All patients provided written informed consent and the study was approved by the institutional review board. Treatment and surveillance were performed in accordance with the NCCN and EAU guidelines on UTUC.^22,23^ Recommendations to forego neoadjuvant chemotherapy, the performance of template-based lymphadenectomy, and administration of adjuvant treatment were made by the multidisciplinary treatment team. Pathologic specimens were reviewed by board-certified genitourinary pathology specialists and classified according to the AJCC Cancer Staging Manual, 8^th^ edition.^24^ Postsurgical surveillance consisted of cross-sectional imaging, cystoscopy, and urine cytology every 3-6mo in the first two years and every 6-12mo thereafter according to the pathologic stage and grade.

Peripheral blood (10 ml) was collected in EDTA-containing tubes (Streck cell-free DNA BCT, La Vista, Nebraska, USA) 1-2 hours prior to surgery. Two-step centrifugation of whole blood was performed at 1600xg for 10 minutes followed by 3200xg for 10 minutes at 10°C. Plasma, buffy coat, and cell pellet were stored at −80°C. From the nephroureterectomy specimen, thick 20 µm sections of formalin-fixed paraffin embedded (FFPE) tumor tissue with ≥50% tumor cellularity were macrodissected for nucleic acid extraction.

### DNA Extraction, Next-Generation Sequencing, and Bioinformatics Analysis

Peripheral blood mononuclear cell (PBMC)-derived germline DNA (gDNA), tumor tissue DNA, and plasma cell-free DNA (cfDNA) were extracted using a combination of established proprietary kits and in-house column-based methods as previously described.^25^ Thirty of 32 patients had adequate preoperative plasma samples for ctDNA extraction. After quality assessment and quantification, up to 250 ng of gDNA, 50-100 ng of tumor DNA, and 5–30 ng of cfDNA were used for next-generation sequencing library preparation, panel-based hybridization (152-gene PredicineCARE© panel; Supplemental Fig. 1)^25^, and enrichment prior to ∼20,000X, 150bp paired-end sequencing on the Illumina NovaSeq 6000 platform (Illumina, San Diego, CA). In parallel, plasma samples were also sequenced using low-pass (1-3X) whole-genome sequencing (WGS).

A proprietary machine learning bioinformatics pipeline (Predicine DeepSEA©) was used to identify single nucleotide variations (SNVs), insertions/deletions (indels), gene-level copy number changes (CNAs), and targeted gene fusions as previously described.^26^ This algorithm incorporates customized probabilistic control for sequencing errors, detects and eliminates mutations potentially resulting from clonal hematopoiesis of indeterminate potential (CHIP), and calls mutations passing validated allele frequency thresholds of 0.25% (or 0.1% for hotspot mutations) in plasma and 5% (or 2% for hotspot mutations) in FFPE. Additionally, germline mutations detected in matched PBMCs or at high frequency in population genomic databases were filtered out. Pathogenicity of mutations was annotated according to the Clinvar data base.^27^ Tumor mutation burden (TMB) was calculated using high-confidence mutations and adjusted for the panel size. In parallel, the low-pass WGS data from plasma cfDNA was used to conduct chromosomal-level copy number analysis using a modified version of the ichorCNA algorithm.^28^ Genome-wide copy number burden (CNB) was calculated as an aggregate score across significant copy number changes detected in 1MB windows throughout the autosomal genome (see Supplemental Methods). Using low-pass WGS data, we tested for evidence of somatic chromosomal arm copy number changes in the plasma cfDNA.

### Outcomes and Statistical Analyses

The primary objective was to investigate the ability of plasma ctDNA to distinguish between muscle-invasive/non-organ confined (MI/NOC) and non-muscle invasive (NMI) UTUC. The predictive performance of ctDNA for preoperative identification of MI/NOC UTUC was summarized across preoperative variant count thresholds by calculating the area under the receiver-operating characteristic curve (AUC). The optimal variant count threshold for best sensitivity and specificity was determined using Youden’s J statistic implemented in the R package pROC.^29^ Based on this method, preoperative ctDNA positivity was defined as the detection of at least two plasma variants, coinciding with other published analyses.^19^ The Kaplan-Meier method was used to estimate survival and Mantel-Cox log-rank testing to assess associations between preoperative ctDNA positivity and clinical outcomes including 1) overall survival (OS, time from surgery until UC-related death) and 2) progression-free survival (PFS, time from surgery until progression to metastatic UC). Differences between NMI and MI/NOC patient groups were tested using the Wilcoxon test for continuous variables and the Fisher Exact test for categorical variables. Univariate analysis of each variable was done using logistic regression, and elastic-net regularization was imposed for multivariate models (R packages glm, caret and glmnet). All tests were conducted in R version 4.1.3.

## RESULTS

### Patient characteristics and UTUC Staging

Overall, 30/32 patients with clinically high-risk UTUC undergoing surgical extirpation had preoperative plasma ctDNA passing quality control (QC). Of the two samples failing QC, one was due to processing error and another due to insufficient DNA yield. Of the remaining 30 patients, median age was 74 years (IQR 67, 77.8) and 21 (70%) were male (Table 1). The tumor was located in the renal pelvis in 10 (33%), the ureter in 9 (30%), and both in 11 (37%). Twenty-seven (90%) patients underwent nephroureterectomy, 3 (10%) segmental ureterectomy, and 12 (40%) concomitant regional lymphadenectomy. Two (7%) patients received topical therapy before definitive surgical extirpation and 6 (20%) received adjuvant/salvage chemotherapy. On surgical pathology, 24 (80%) were high grade and 13 (43%) were MI UTUC (≥pT2). Six (20%) patients were found to harbor nodal metastases. Interestingly, one patient with pT1 HG ureteral tumor was found to have occult nodal metastasis, resulting in 14/30 (47%) patients with MI/NOC UTUC (Fig. 1).

**Table 1.**
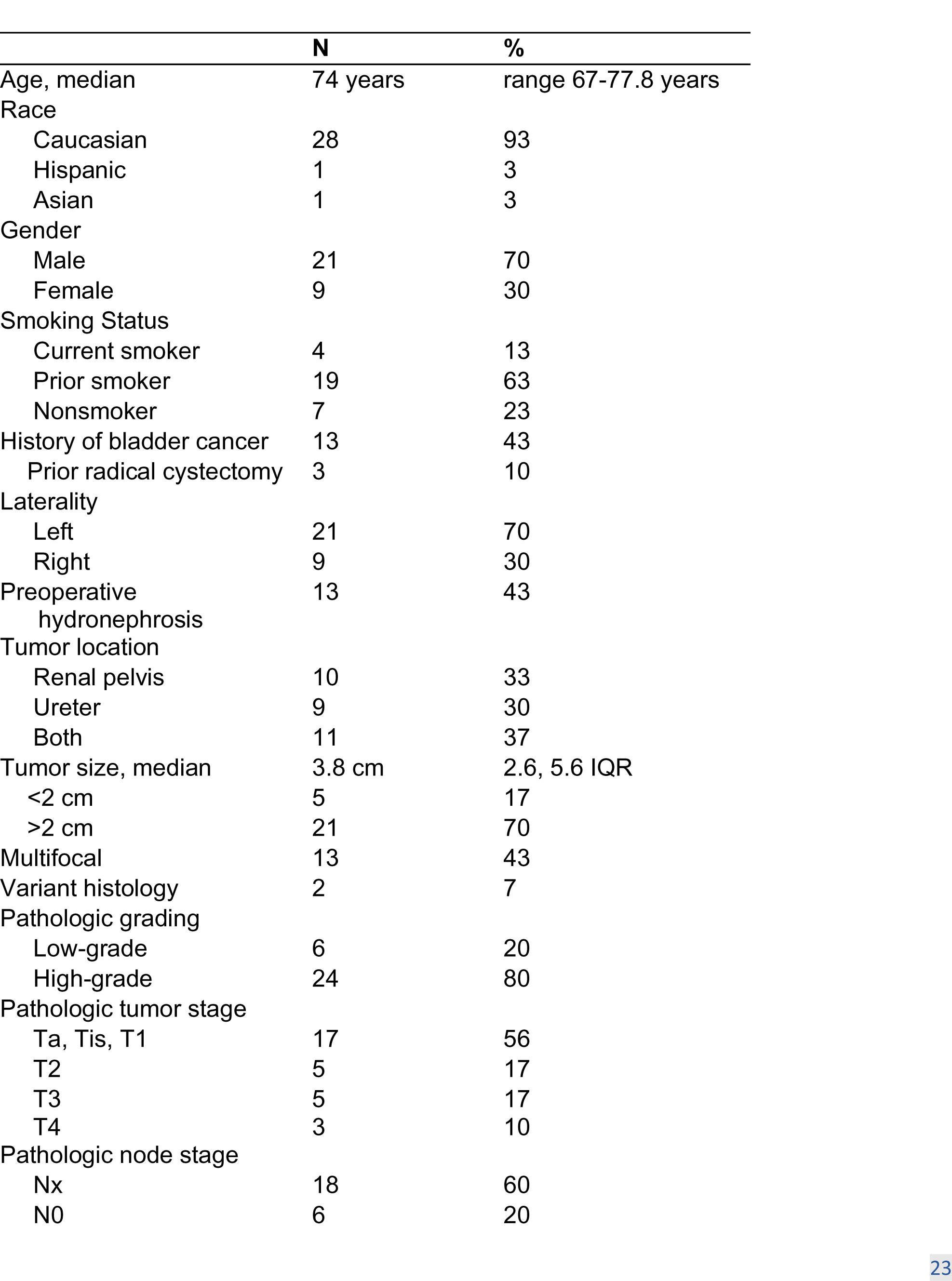
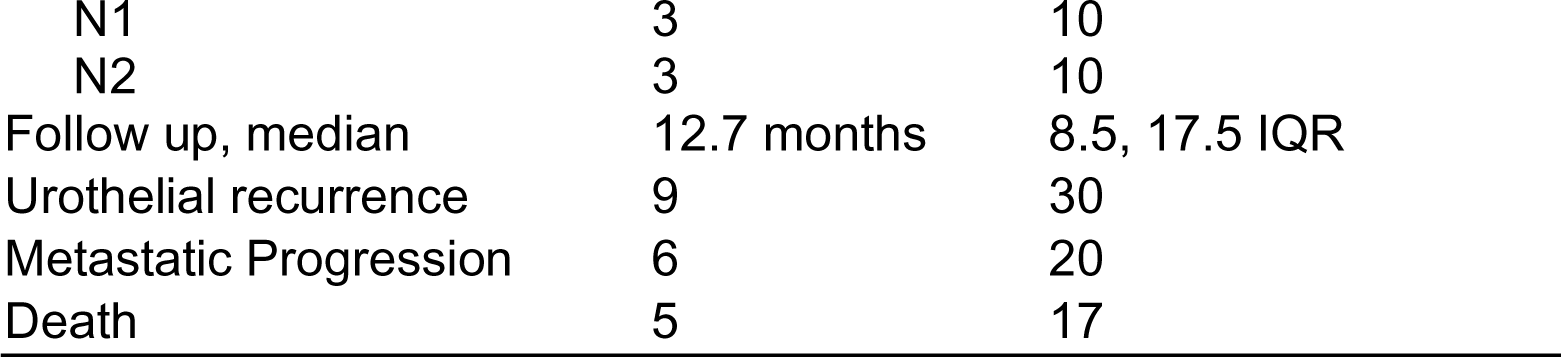
Clinicopathologic and treatment characteristics (N=30)

|  | N | % |
| --- | --- | --- |
| Age, median | 74 years | range 67-77.8 years |
| Race |  |  |
| Caucasian | 28 | 93 |
| Hispanic | 1 | 3 |
| Asian | 1 | 3 |
| Gender |  |  |
| Male | 21 | 70 |
| Female | 9 | 30 |
| Smoking Status |  |  |
| Current smoker | 4 | 13 |
| Prior smoker | 19 | 63 |
| Nonsmoker | 7 | 23 |
| History of bladder cancer | 13 | 43 |
| Prior radical cystectomy | 3 | 10 |
| Laterality |  |  |
| Left | 21 | 70 |
| Right | 9 | 30 |
| Preoperative hydronephrosis | 13 | 43 |
| Tumor location |  |  |
| Renal pelvis | 10 | 33 |
| Ureter | 9 | 30 |
| Both | 11 | 37 |
| Tumor size, median | 3.8 cm | 2.6, 5.6 IQR |
| <2 cm | 5 | 17 |
| >2 cm | 21 | 70 |
| Multifocal | 13 | 43 |
| Variant histology | 2 | 7 |
| Pathologic grading |  |  |
| Low-grade | 6 | 20 |
| High-grade | 24 | 80 |
| Pathologic tumor stage |  |  |
| Ta, Tis, T1 | 17 | 56 |
| T2 | 5 | 17 |
| T3 | 5 | 17 |
| T4 | 3 | 10 |
| Pathologic node stage |  |  |
| Nx | 18 | 60 |
| N0 | 6 | 20 |
| N1 | 3 | 10 |
| N2 | 3 | 10 |
| Follow up, median | 12.7 months | 8.5, 17.5 IQR |
| Urothelial recurrence | 9 | 30 |
| Metastatic Progression | 6 | 20 |
| Death | 5 | 17 |

**Figure 1.**
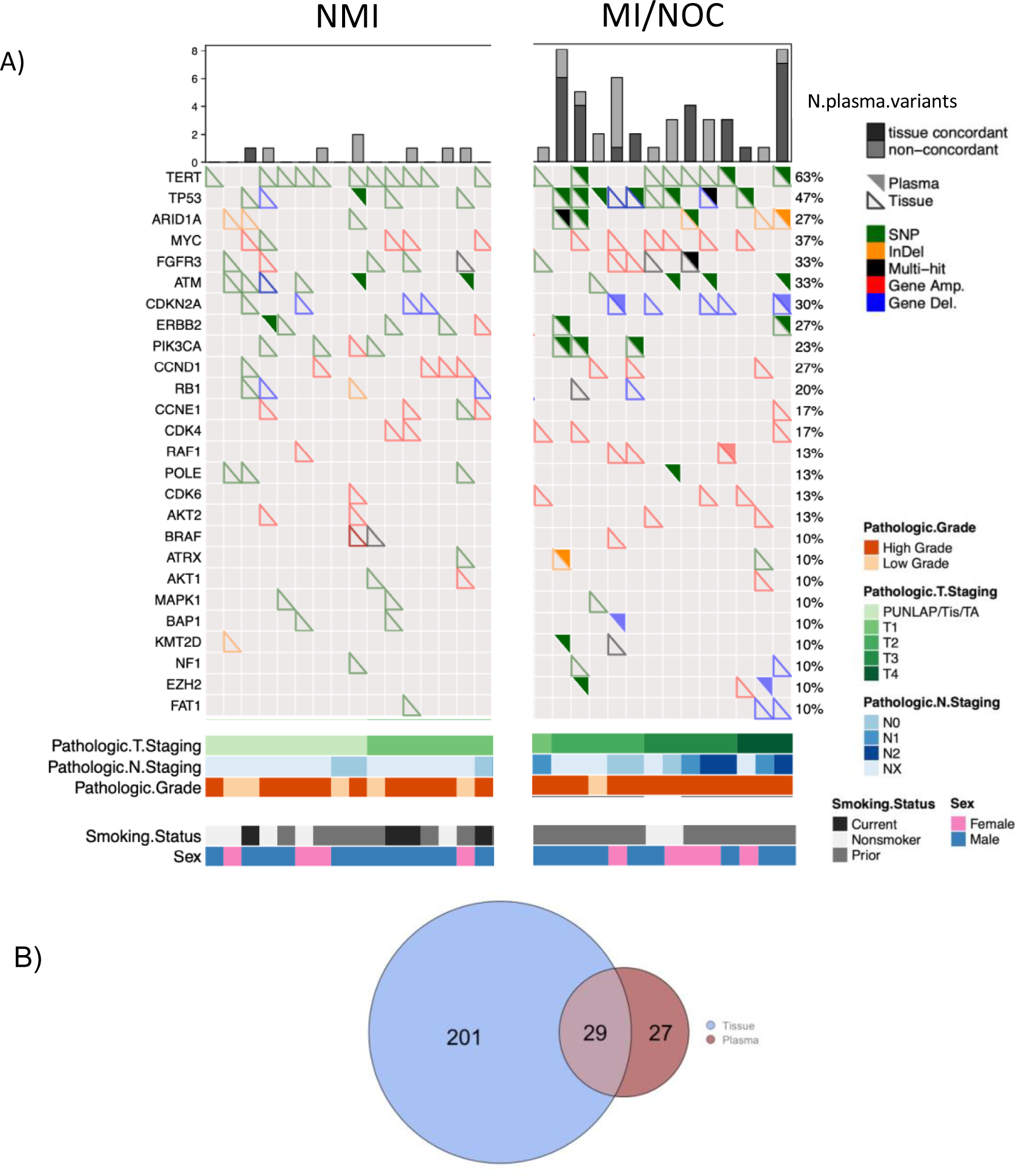
Summary of molecular alteration profiling, histopathologic, and clinical characteristics for 16 NMI and 14 MI/NOC UTUC patients with paired tumor tissue and preoperative plasma cfDNA profiles ordered by advancing pathologic T stage. Each column represents one patient. Upper filled triangles represent plasma mutations and lower open triangles represent tumor tissue mutations. B) Venn diagram showing overall mutational concordance between tumor tissue and plasma cfDNA.

### Somatic mutations and CNAs

The 152-gene PredicineCARE™ panel covers 81.2% of the commonly altered genes (≥10% incidence) in UTUC.^30^ To investigate the concordance between plasma and tumor tissue-derived DNA, targeted sequencing was performed on matching plasma and surgical UTUC samples. Overall, molecular alterations (MA) including SNVs/indels (66%) and gene-level CNAs (34%) were detected in 29 out of 30 (97%) tumor samples, spanning 75 of the 152 paneled-genes (Fig. 1a). In addition, 1 *FGFR3*-*TACC3* fusion^31^ was found. Of the SNVs and indels, 29% were classified as pathogenic.^27^ Each tumor contained a median of 6 (range 0-18) MAs and a mean TMB of 8.8 mutations/Mb (range 0-35.1), similar to levels previously reported.^30,32^ One (3.3%) hypermutated tumor was found within our cohort (Supplemental Fig. 2), consistent with the 5.5% incidence described by Fujii et al.^30^ Interestingly, this patient did not have germline mismatch repair gene alterations or prior cancer history. The most common tumor derived variants included *TERT* promoter (63%), *TP53* (40%), *MYC* (37%), *FGFR3* (33%), and *CDKN2A* (30%) (Fig. 2b), in line with previous reports.^30,32^ Although there was no significant difference in the total number of variants detected between NMI vs MI/NOC tumor tissue (8.3 vs 7, p=0.5, Fig. 2a), important distinctions exist at the gene level. Similar to previous reports^30,32^, *TP53* (57% vs. 25%, p=0.1) was more commonly mutated in MI/NOC UTUC though this difference did not reach statistical significance. In contrast, a similar prevalence of *FGFR3* (31% vs. 31%) and *TERT* promoter (69% vs. 57%) alterations were found in NMI and MI/NOC tumors (Fig. 2b).

**Figure 2.**
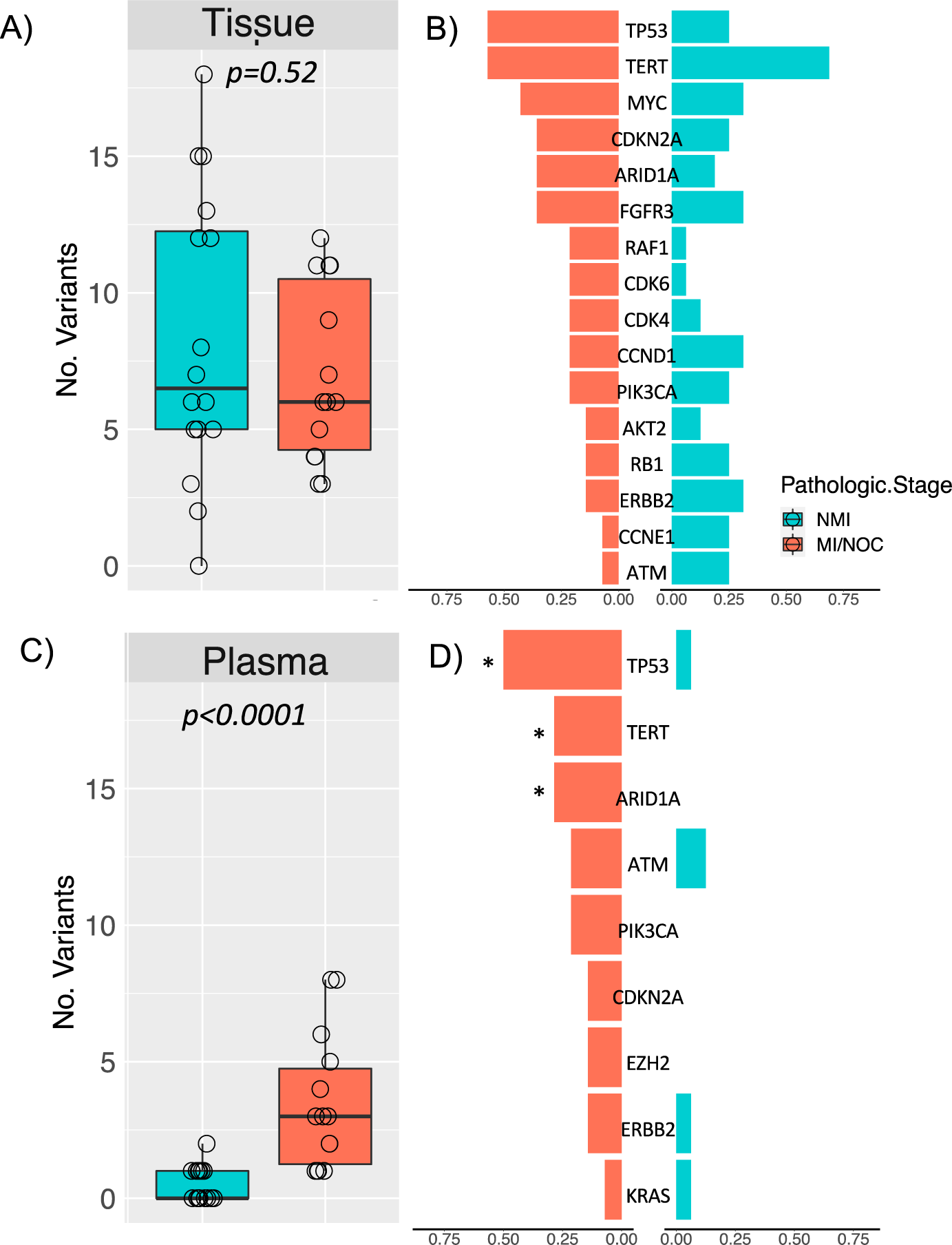
Numbers of molecular alterations and their frequencies found in the tumor tissue (A-B) and plasma cfDNA (C-D). There was a significantly higher number of alterations observed in the plasma of Ml/NOC vs. NMI patients (3.4 vs. 0.5, p<0.0001), but not in the tumor tissue (7.0 vs 8.3, p=0.52). No gene was significantly more frequently altered in Ml/NOC compared to NMI tumor tissue. However, alterations were more frequently found in TP53, TERT, and ARID1A in the plasma from Ml/NOC patients (*). Tissue molecular alterations found in 3 or more patients and plasma alterations in 1 or more are shown.

At least one MA was detected within the cfDNA from 21/30 (70%) preoperative plasma samples, with each patient carrying a median of 1 ctDNA variant (range 0-8) (Fig. 1a). The most commonly detected alterations in the plasma cfDNA were *TP53* (27%), *ATM* (17%), and *ARID1A* (13%) (Fig. 2d). Overall, 52% of the detected plasma ctDNA variants corroborated alterations detected in the matching tumor samples. On the other hand, 88% of the tumor variants were not detected within the plasma (Fig. 1b). In particular, alterations in *TP53*, *ARID1A*, and *PIK3CA* were frequently detected concomitantly in paired plasma and tissue samples. Importantly, a significantly higher number of plasma variants were detected within the plasma from MI/NOC (mean 3.4, range 1-8) vs. NMI UTUC (mean 0.5, range 0-2, p<0.0001) (Fig. 2c).

### Clinical Utility of Preoperative ctDNA Detection and Staging

The detection of ctDNA has previously been utilized in the preoperative setting to estimate tumor burden and refine prognosis in patients with stage III cutaneous melanoma.^33^ Similarly, we evaluated the utility of preoperative ctDNA to predict clinical staging in our cohort. Following summarization of predictive performance for optimal sensitivity and specificity of preoperative identification of MI/NOC UTUC across variant count thresholds, the presence of pre-surgical ctDNA was defined as the detection of at least two panel-based plasma MAs. Detection of ctDNA at this threshold was strongly predictive of MI/NOC UTUC at the time of surgery, achieving a sensitivity of 71% and specificity of 94% for detecting MI/NOC UTUC stage with an AUC of 0.92 (0.85-1.0, 95% CI, Supplemental Fig. 3). From 14 MI/NOC patients, four false negatives occurred, in patients with HG-UTUC staged pT1N1, pT3Nx, pT4Nx, and pT4N1. The one false positive occurred in a patient with multifocal HG pTaN0 with a large 6.1 cm renal pelvic tumor.

Moreover, the presence of ctDNA preoperatively was strongly prognostic. At a median follow-up of 12.7 months (Range 8.5-17.5 months), 6 (20%) patients suffered progression to metastases and 5 (17%) disease-specific death. Of the 11 patients with detectable ctDNA (with at least two panel-based plasma variants), 5 (45%) were found to have disease progression, 3 (27%) died of UTUC, and 1 (9%) died of unknown causes (Fig. 3). In contrast, 1 of 19 (5%) patients with negative ctDNA suffered disease progression and died (Fig. 3). Patients with detectable preoperative ctDNA had significantly shorter progression-free (one-year PFS 69% vs 100%, p<0.001) (Fig. 4a) and overall survival (56% vs 100%, p<0.02) (Fig. 4b). ctDNA positivity predicted both shorter PFS (HR=20.4, 95% CI [2.4, 174.3], p<0.0001) and OS (HR=9.3, 95% CI [1-84], p=0.02). In contrast, preoperative ctDNA positivity was not significantly associated with urothelial recurrence-free survival (one-year RFS 57% vs 59%, p=0.1).

**Figure 3.**
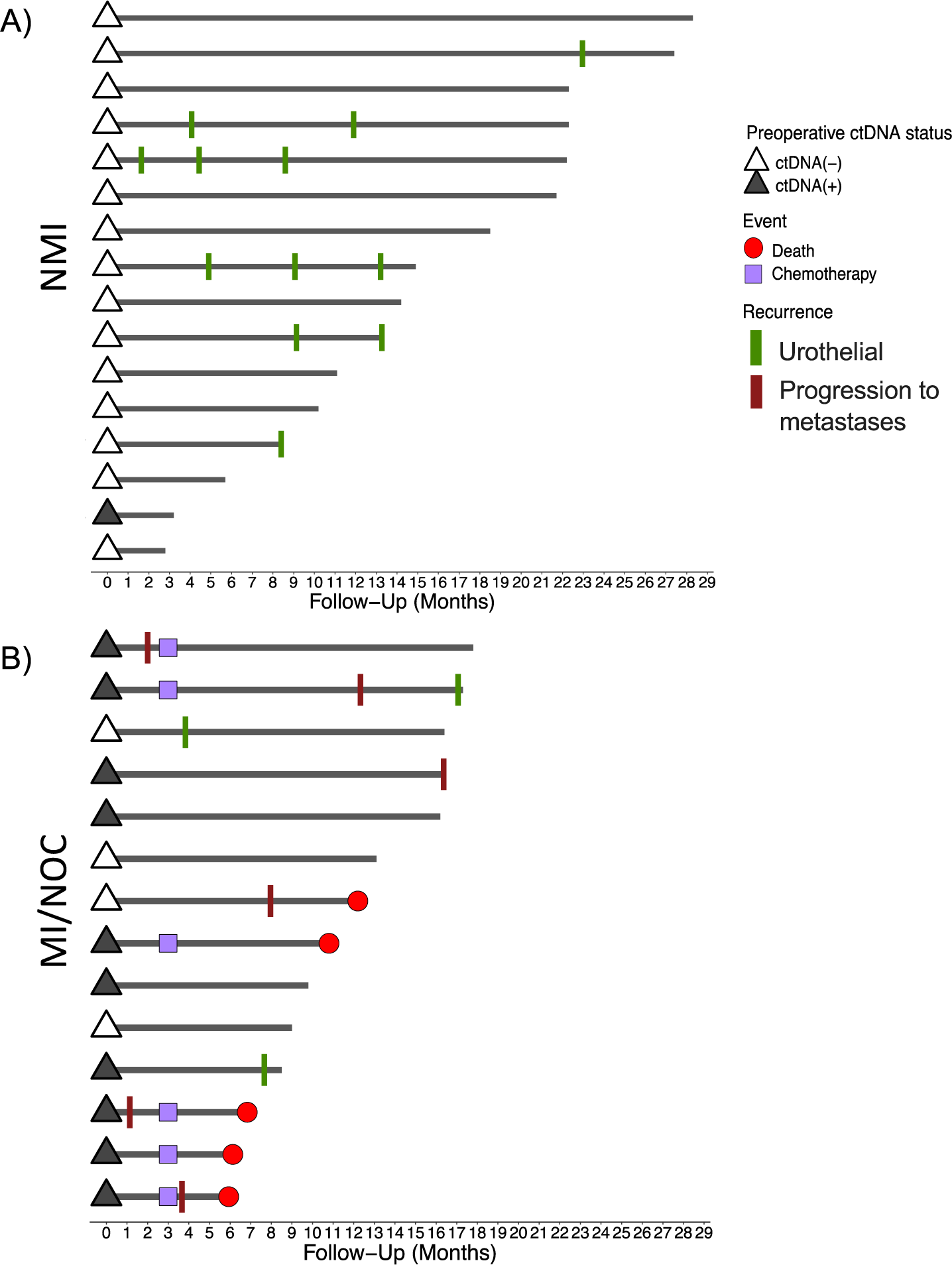
An event chart showing patients with A) NMI and B) Ml/NOC UTUC. Preoperative ctDNA positivity (</= 2 plasma variants detected) is shown as open and filled triangles at time of surgery. Urothelial recurrences and progression to metastases are shown during follow up along with adjuvant chemotherapy treatment.

**Figure 4.**
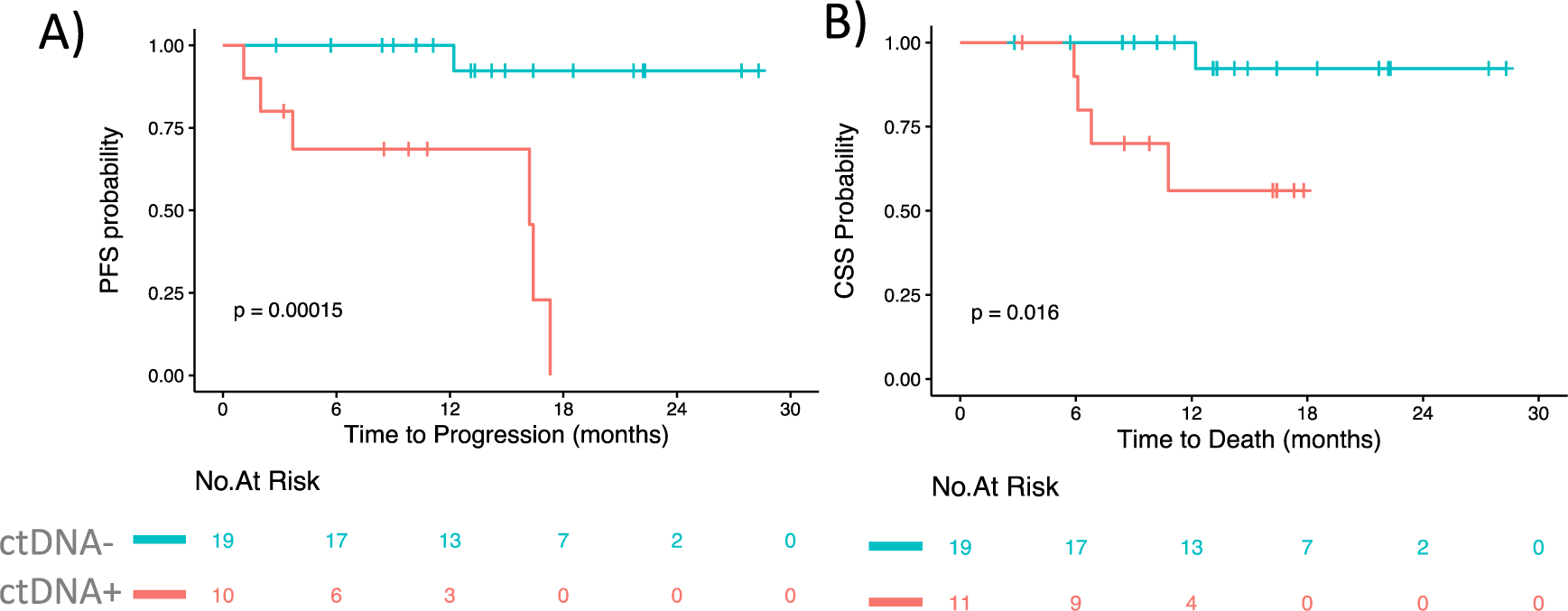
Prognostic value of ctDNA detection. A) Progression-free and B) overall survival were significantly prolonged for those patients who were ctDNA positive at the time of extirpative surgery. One-year PFS was 69% for patients with positive preoperative ctDNA compared to 100% for patients with negative ctDNA (p<0.001). Similarly, one-year OS was 56% compared to 100% (p=0.016) at a median 12.7 months of follow up.

### Utility of Copy Number Changes and Staging

Previous studies have demonstrated a dichotomy between UTUCs with highly complex karyotypes containing frequent focal CNAs, aneuploidy, and chromothripsis compared to those with simple arm-level aberrations most commonly involving chromosome 1q, 3, 8, and 9.^30^ Furthermore, those with complex CNAs were frequently found to have *TP53* alterations, and more likely to be staged as muscle-invasive and exhibit aggressive phenotypes.^30^ We hypothesized that high CNB correlates with pathologic staging and can be used to complement ctDNA positivity to improve the prediction model for MI/NOC UTUC. Evidence of somatic chromosomal arm gain and/or loss was found in all tumors (Supplemental Table 1). As previously reported^30^, *TP53*-altered tumors had numerically higher number of CNAs than other subtypes (3.5 vs. 2.1, p=0.09, Supplemental Fig. 4). Overall, there was a marginal difference in preoperative plasma CNB score (4.5 vs. 5.2, p=0.06) between NMI and MI/NOC UTUC patients (Supplemental Fig. 5A). Imposing a threshold of plasma CNB score >6.5 to confirm MI/NOC UTUC when at least two plasma variants were detected increased the sensitivity of prediction to 79%, without compromising the specificity (Supplemental Fig. 5B).

## DISCUSSION

The inability to accurately stage tumors prior to surgical extirpation has severely hampered customization of treatment for patients with high risk UTUC. As evidenced herein, despite adherence to clinical guidelines, 30% of the patients suffered rapid disease progression and/or cancer-specific death within two years following surgery with curative intent. With emerging evidence supporting the use of neoadjuvant chemotherapy, clinical tools to enhance the identification of potential beneficiaries with invasive, micrometastatic disease prior to surgery are critically needed. The inherent loss of renal function from surgical extirpation renders a subset of patients ineligible for adjuvant cisplatin-based chemotherapy, which makes this unmet need even more dire.^11^ Against this backdrop, our study provides encouraging evidence that plasma ctDNA collected at diagnosis estimates tumor burden and can be leveraged to distinguish between patients with MI/NOC from those with NMI UTUC. Equally important, ctDNA was strongly prognostic for disease progression and death following surgery, making it a promising biomarker for selecting patients to undergo chemotherapy in the neoadjuvant setting. These results corroborate what has previously been described in muscle invasive bladder cancer.^19,34^

When treating high risk UTUC, more emphasis should be placed on minimizing missed opportunities to provide life-prolonging systemic treatment to patients with MI/NOC UTUC undergoing extirpative surgery. To that end, the sensitivity of preoperative ctDNA to define MI/NOC UTUC reached 79% in our model based on the detection of at least 2 panel-based plasma variants and a minimal threshold of plasma CNB score of 6.5. Although needing validation, this level of sensitivity represents a clear improvement over the more modest sensitivities between 42 – 48% achieved using available clinical nomograms.^7,8^

On top of its clinical relevance, the 152-gene PredicineCARE™ ctDNA platform provided ease of clinical application and high genomic fidelity. Of the 31 plasma samples appropriately processed, only 1 failed to yield sufficient cfDNA for analysis. From the analysis of tumor tissue samples, MAs were detected in all but one of the samples (97%), validating the broad coverage of the frequently altered genes in UTUC.^30,32^ Furthermore, the tissue genomic landscape as detected by the panel (including frequent alterations in *TERT* promoter, *TP53, FGFR3* and *CDKN2A*) resembled that described in previous datasets.^30,32^

Due to the rare incidence of UTUC, scant data exists on the application of ctDNA in the localized UTUC setting. Using a 73-gene panel, Agarwal et al^14^ defined ctDNA as one or more tumor-derived MAs and reported detection of ctDNA in 95% of 75 metastatic UTUC patients from 13 academic institutions, with an average of 6.8 MAs per patient. The higher plasma MA rates may reflect higher disease burden in patients with metastatic disease, though sporadic UTUC has also been shown to have a lower mutational burden than urothelial carcinoma of the bladder.^35^ Similar to our study, the most frequently encountered plasma MAs were *TP53* (51%), *PIK3CA* (23%), *ARID1A* (20%), and *TERT* (17%), albeit at higher detection frequencies. Likewise, frequent chromosomal arm-level gains and losses in addition to focal CNAs at the 9p24.3 region (CD274, JAK2, and PDCD1LG2) and 1q21.3-1q23.3 (PVRL4) were detected in our study as previously described in UTUC tissue samples.^30^ These genome-wide measures of plasma cfDNA CNB were also associated with advanced staging (5.2 vs. 4.5, p=0.06). Adding CNB as a secondary predictor of MI/NOC UTUC to our model increased the sensitivity from 71% to 79% (Supplemental Fig. 5B). Our study represents the first investigation at scale of the ctDNA landscape in localized, high-risk UTUC and its concordance with somatic mutations found from matched tumor specimens. The biological implications extending from this study requires further investigation and is beyond the scope of the current report.

Our study was certainly not without limitations. First, the small sample size reflects the rarity of the disease. Even in a high-volume tertiary referral center, this study spanned 1.5 years. Future multi-institutional validation efforts are required to prove the clinical value of this test. Secondly, rather than testing for tissue-informed mutations in a bespoke manner, a paneled genomic test was chosen. This strategy specifically addresses the clinical challenge of obtaining a small quantity of biopsy samples during the diagnosis of UTUC (limiting genomic sequencing) as well as the urgency to reach a treatment decision. Additionally, diagnostic ureteroscopic biopsy may also result in other untoward consequences, such as increased intravesical tumor recurrence.^36^

Although the PredicineCARE™ platform was not specifically designed to diagnose UTUC, it covers 81.5% of the common UTUC MAs. Furthermore, the predictive power of the test was not improved by limiting ctDNA positivity to only those alterations previously associated with UTUC (Supplemental Fig. 6) nor with multivariate modeling incorporating clinical characteristics (Supplemental Fig. 7). Lastly, as paneled sequencing is not ideal for the detection of minimal residual disease, alternative bespoke strategies based on MAs obtained from the surgical tissue sample may yield higher clinical impact for the detection of minimal residual disease following surgery. Nevertheless, the strong prognostic power demonstrated in our study represents a major step towards customization of treatment.

## CONCLUSION

In this prospective observational study, we demonstrate the clinical utility of ctDNA managing high risk, localized UTUC. Preoperative ctDNA positivity based on the detection of at least 2 plasma variants and a minimal threshold of CNB score of 6.5 was highly predictive of MI/NOC UTUC staging and strongly prognostic of progression and overall survival. Preoperative ctDNA analysis is feasible and may be used to select high risk UTUC patients benefiting from neoadjuvant chemotherapy.

## Data Availability

All data produced in the present study are available upon reasonable request to the authors

## Acknowledgments

This research was supported by the H Lee Moffitt Cancer Center & Research Institute; Predicine, Inc; and the American Cancer Society-Institutional Research Grant from January 1-December 31, 2021. EAS and ACS were the recipients of support from the Junior Scientist Research Partnership award from Moffitt Cancer Center and Research Institute.

## ABBREVIATIONS

AUC: Area under a receiver-operating curve
cfDNA: Cell-free DNA
CNA: Copy number alterations
CNB: Copy number burden
ctDNA: Circulating tumor DNA
FFPE: Formalin-fixed paraffin-embedded
GA: Genomic alterations
gDNA: Germline DNA
indel: Small insertion or deletion
MA: Molecular alteration
MI/NOC: Muscle-invasive/non-organ confined UTUC
NAC: Neoadjuvant chemotherapy
NMI: Non-muscle-invasive UTUC
PBMC: Peripheral blood mononuclear cell
PFS: Progression-free survival
RNU: Radical nephroureterectomy
SNV: Single nucleotide variation
UTUC: Upper tract urothelial carcinoma
WGS: Whole genome sequencing

---

**Supplemental Figure 1.**
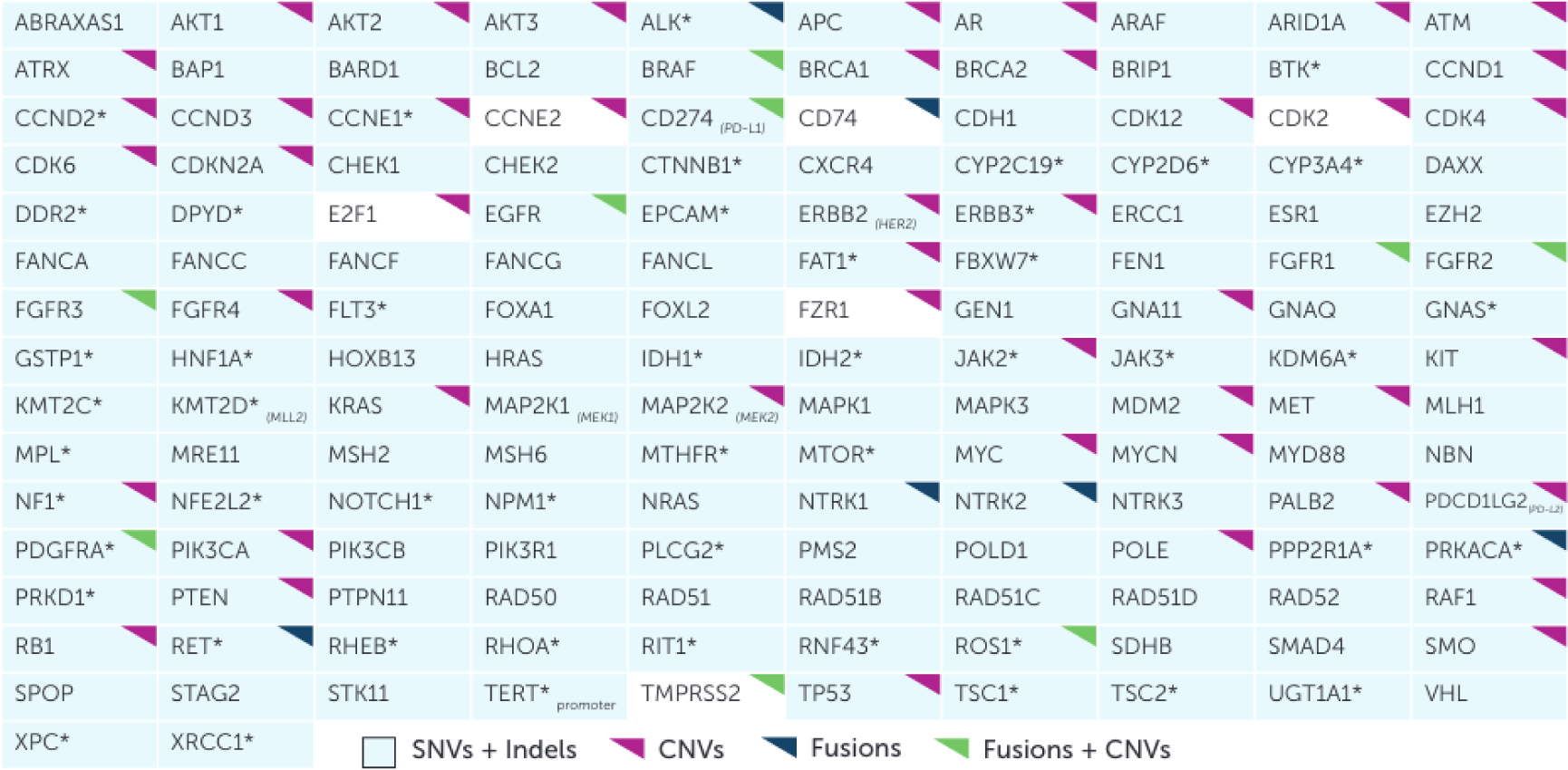
Genesincluded in the PredicineCARE™assay. The panel interrogates 152 genes, including 103 genes with complete exonic coverage and 49 genes with select exonic coverage (indicated with *). PredicineCARE™ also includes parallel low-pass WGS sequencing used to generate CNB Score and analysis of large-scale chromosomal amplificationsand deletions.

**Supplemental Figure 2.**
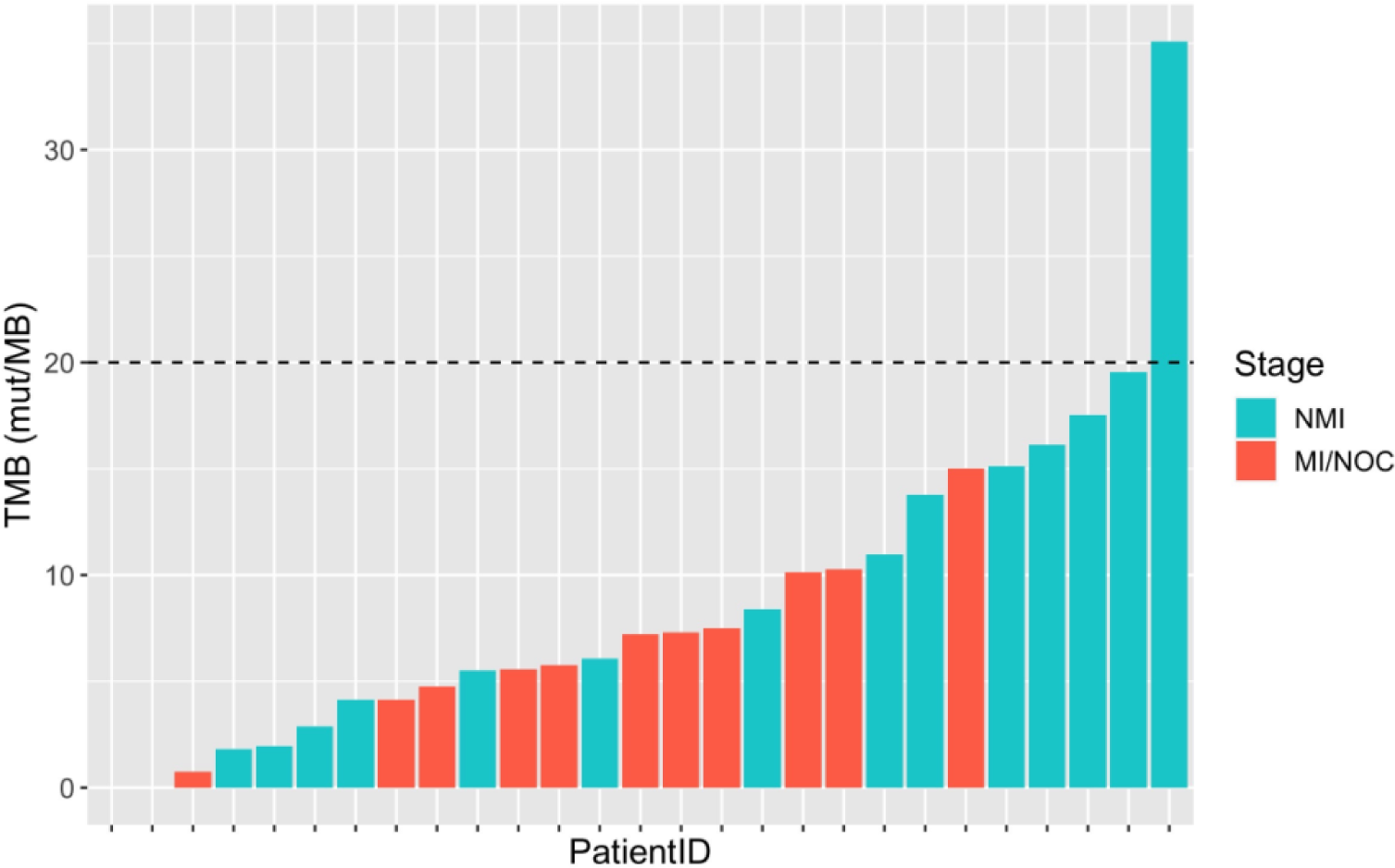
UTUC tumor tissue-based TMB scores for patients. The highest scoring patient with TMB >20 mut./Mb can be classified as hypermutated.

**Supplemental Figure 3.**
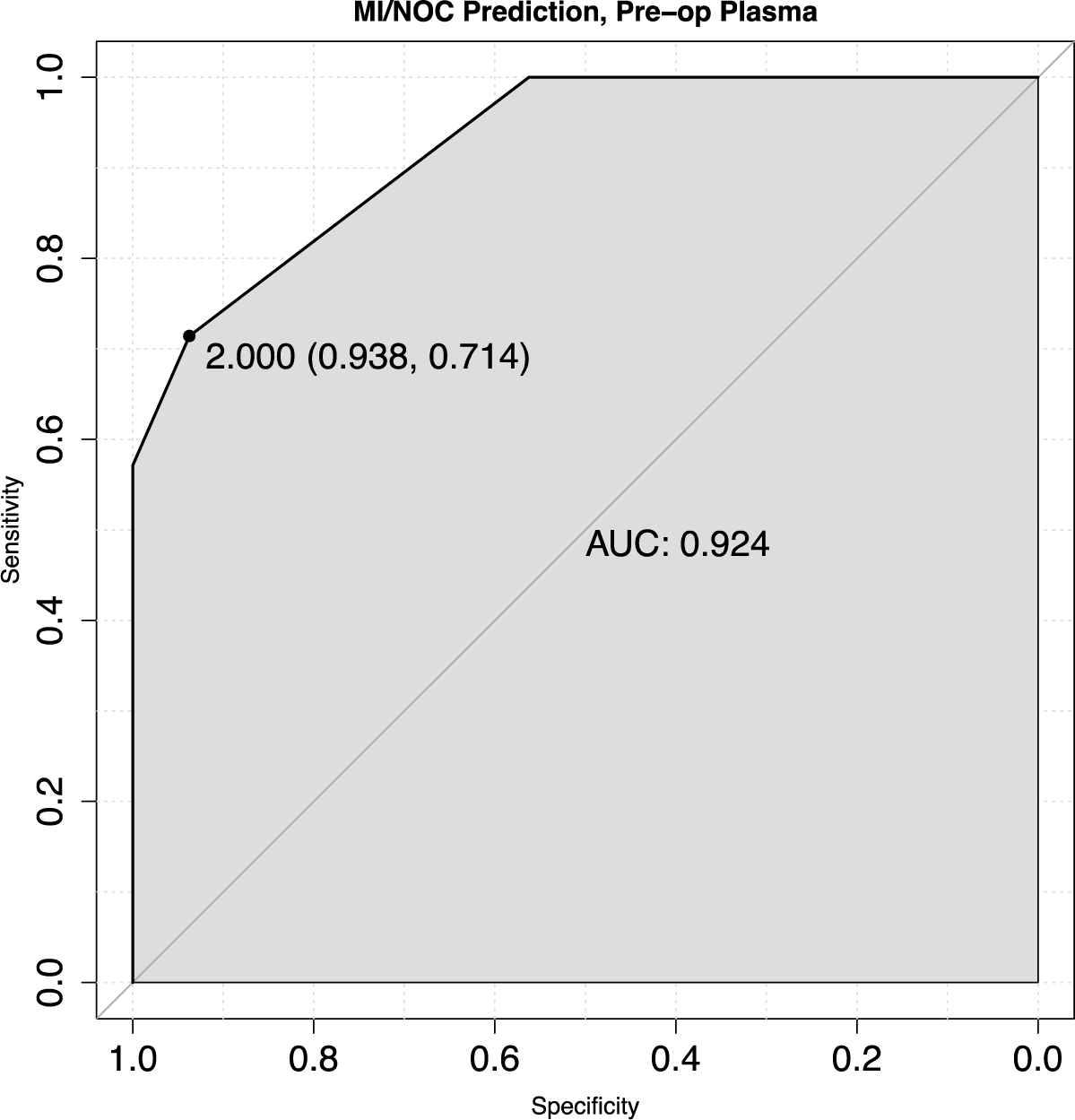
Receiver-operating curve for prediction of Ml/NOC UTUC from preoperative plasma cfDNA variant count (including SNVs, indels, and CNV).

**Supplemental Figure 4.**
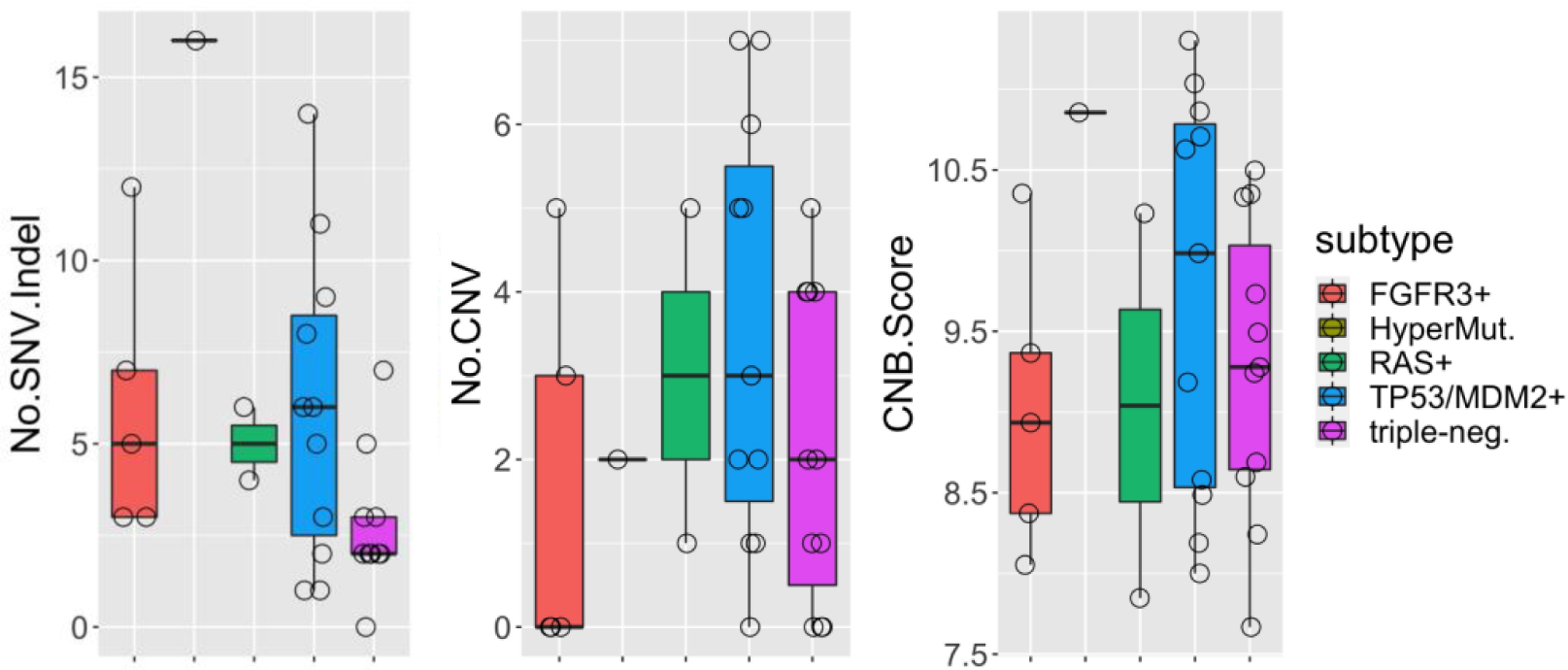
SNV counts, CNV counts, and CNB scores from tumor tissue for patients from this study classified according to mutational subtypes defined by Fujii et al. 2021. Patients with the TP53+ mutational subtype have a significantly higher number of gene-level CNVs (1.9 vs. 3.7, *p =0.03*) and a marginally higher number of SNV/indels (3.8 vs. 5.6, *p=0.13*) and genome-wide copy-number burden (9.1 vs. 9.8, *p =0.09*). The hypermutated subtype sample is also positive for TP53 mutation.

**Supplemental Figure 5.**
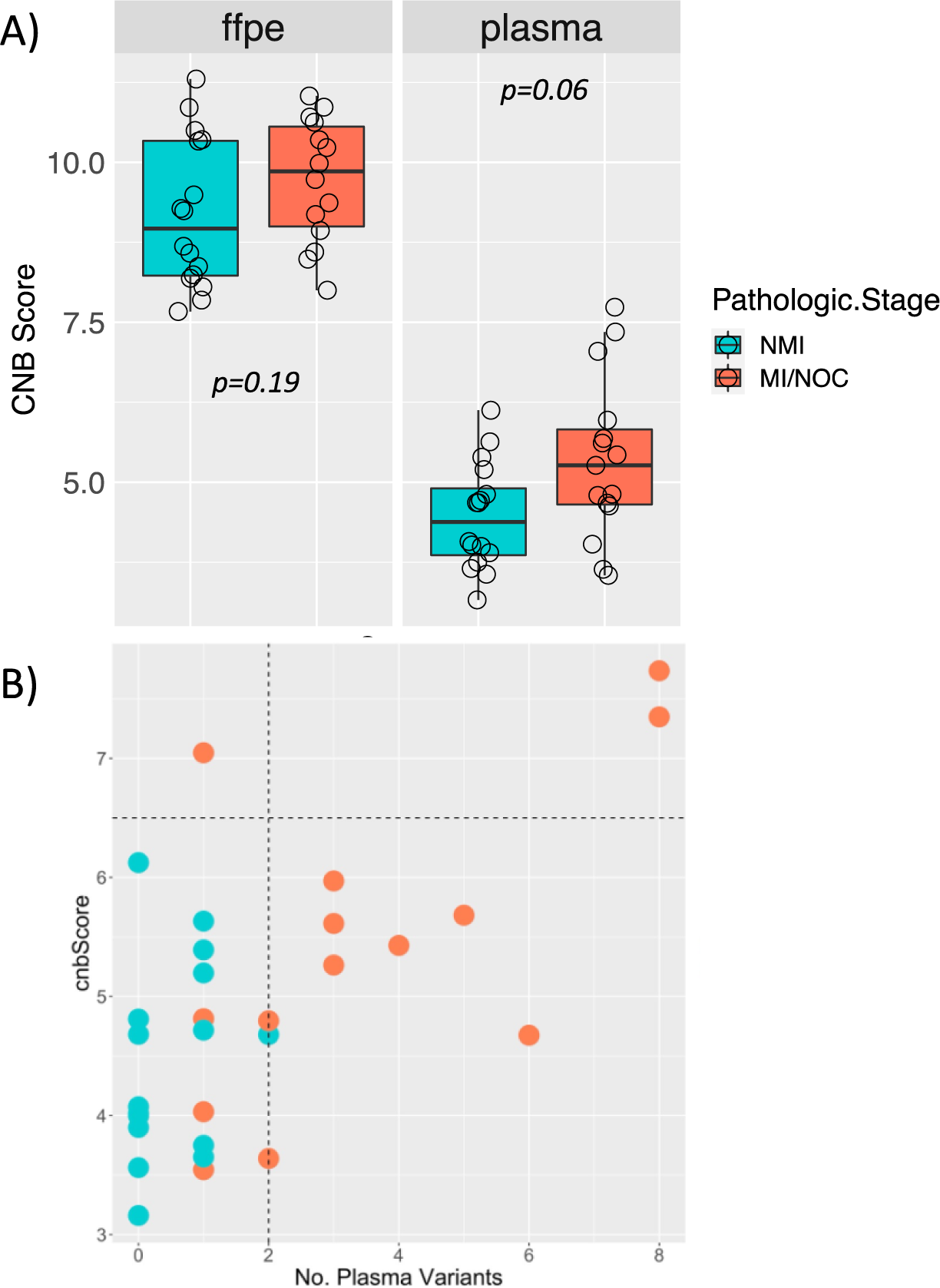
A) CNB score of tumor tissues and preoperative plasma cfDNA for NMI and Ml/NOC patients. B) Plasma CNB score vs. number of variants observed for each patient. Setting a threshold of plasma CNB Score >6.5 (horizontal dotted line) to confirm Ml/NOC disease for patients with 2:2 observed plasma variants (vertical dotted line) adds one additional true positive call (upper left quadrant) without any reduction in specificity. Overall sensitivity of this stepwise method is 79% at 94% specificity.

**Supplemental Figure 6.**
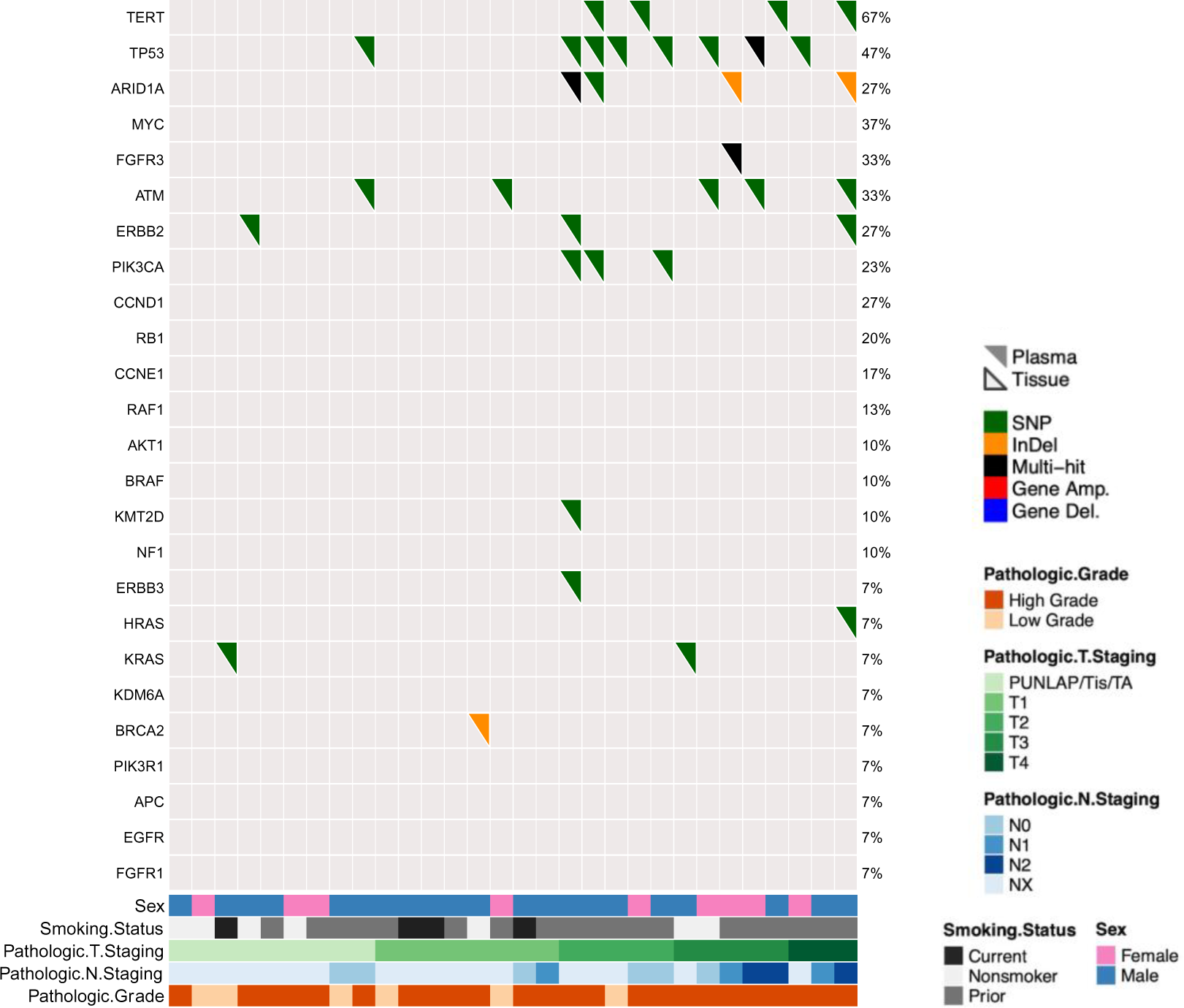
Oncoprint showing tissue and plasma mutations observed for the 25 most frequently mutated genes out of a list of 64 genes observed as mutated in UTUC in other studies. Each column represents one patient. Upper solid triangles in the grid are mutations in plasma ctDNA and lower open triangles are mutations in paired tumor tissue. Using this UTUC-specific mutational profile, ctDNA detected MI/NOC staging with a preserved specificity of 94% but lower sensitivity of 57% (AUC 0.85, 95% CI 0.71-98).

**Supplemental Figure 7.**
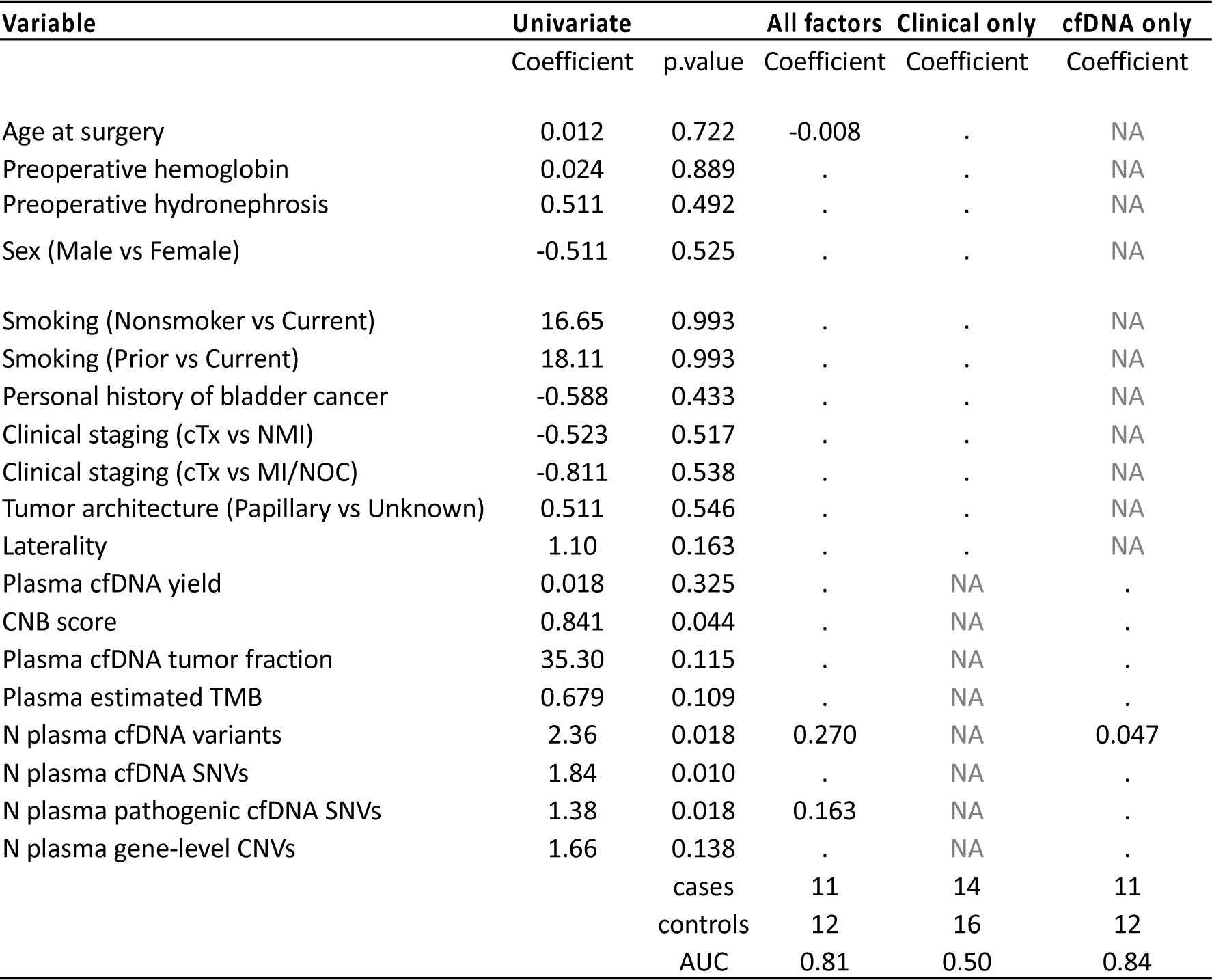
Univariate regression modeling and elastic-net regularization demonstrates that multivariate models did not improve predictive power compared to the simple use of the number of plasma MAs for prediction of MI/NOC UTUC (AUC=0.92). Coefficients that were reduced to zero in the regularized models are shown as a dot. Variables that were not included in each model are shown as NA. For each multivariate model, only patients with complete measurements for all factors were included.

## SUPPLEMENTARY METHODS

### cfDNA and genomic DNA extraction

Whole blood samples were shipped to Predicine on ice and later centrifuged in the lab. Circulating cell-free DNA was extracted from the plasma fraction by QIAamp circulating nucleic acid kit. Quantity and quality of the purified cfDNA was checked using Qubit fluorimeter and Bioanalyzer 2100. For samples with severe genomic contamination from peripheral blood cells, a bead-based size selection was performed to remove large genomic fragments. Genomic DNA (gDNA) was also extracted from the matched buffy coat fraction (PBMCs) from each blood sample and matched patient FFPE tumor sample. Up to 250ng genomic DNA from PBMC and up to 100 ng gDNA was then enzymatically fragmented and purified.

### Library preparation, capture and sequencing

Five to thirty nanograms of extracted cfDNA were used for plasma cfDNA library construction including end-repair, dA-tailing, and ligation of unique molecular identifiers (UMIs) and sequencing adapters. Ligated fragments were amplified via PCR. The amplified DNA libraries were then further checked using Bioanalyzer 2100 and samples with sufficient yield were used for hybrid capture. When available, a subsample of material was reserved for later low-pass WGS.

Library capture was conducted using Biotin labeled DNA probes. In brief, the library was hybridized overnight with the PredicineCARE panel and paramagnetic beads. The unbound fragments were washed away, and the enriched fragments were amplified via PCR. The purified product was checked using a Bioanalyzer 2100 and then loaded onto the Illumina NovaSeq 6000 for NGS sequencing with paired-end 2x150bp sequencing kits.

### Analyses of NGS data

Data was analyzed using the Predicine DeepSea analysis pipeline, which starts from the raw sequencing data (BCL files) and outputs final variant calls. Briefly, the pipeline first does adapter trim, barcode checking, and correction followed by paired FASTQ read alignment to human reference genome build hg19 using the BWA. Candidate variants, consisting of point mutations, small insertions and deletions, are identified across the targeted regions covered in the panel.

### Variant Calling and Annotation

Candidate variants with low base quality, mapping scores, and other quality metrics are filtered. Sequencing and PCR errors are also corrected using the algorithm described in (Newman et al. 2016). In general, a variant identified in cfDNA was considered a somatic mutation only when (i) at least three distinct fragments (at least one of them double-stranded) contained the mutation; and (ii) the mutant allele frequency was higher than 0.25%, or 0.1% for hotspot mutations; and (iii) the ctDNA variant containing fragments are significantly over-represented in comparison with the matched PBMC sample using a fisher-exact test (p-value < 0.01 and odds-ratio > 3). Non-hotspot variants with high variant frequency (> 30%) were considered as suspicious germline variants. For FFPE samples we included variants with >5% MAF, >2% MAF in hotspots. Candidate somatic mutations were annotated for their effect on protein coding genes as well as probable pathogenicity using the ClinVar database and annotation tool Varsome . Intronic and silent changes were excluded from our analyses, while missense mutations, nonsense mutations, frameshifts, or splice site alterations were retained. We also excluded common germline variants annotated in the 1000 genomes, ExAC, gnomAD and KAVIAR databases with population allele frequency >0.5%. Finally, we excluded hematopoietic expansion (CHIP) related variants that have been previously described, including those in DNMT3A, ASXL1, TET2.

### Copy Number Alterations and Copy Number Burden

Copy number variation was first estimated at the gene level using the NGS panel data. The in-house pipeline calculates the on-target unique fragment coverage based on consensus bam files, which are first corrected for GC bias and then adjusted for probe-level bias (estimated from a pooled reference). Each adjusted coverage profile is self-normalized (assuming diploid status of each sample) and then compared against correspondingly adjusted coverages from a group of normal reference samples to estimate the significance of each copy number variant. To call an amplification or deletion of a gene, we required the absolute z-score and copy number change to pass minimum thresholds.

We measured genome-wide copy-number burden with PredicineCNB™(ref).The ichorCNA algorithm [Adalsteinsson VA, et al.] was applied to GC and mappability-normalized reads to estimate plasma and tissue copy number variations using a hidden Markov model (HMM). Firstly, we measured segment level (1MB) copy number deviation as the log2 ratio of the normalized reads between a sample and normal plasma background (or used a normal gDNA background for tissue CNB), then we quantified arm-level CNV deviation as the average of segment CNVs across each chromosome arm. Our method also takes into account local cfDNA fragment-size distributions. Finally, we calculated sample-level copy number burden (CNB score) as the sum of absolute zscore of arm-level CNV deviation, where higher CNB score indicates greater absolute CNV abnormality compared with normal background.

### Gene Fusions

DNA rearrangement was detected by identifying the alignment break points based on the BAM files before consensus filtering. Suspicious alignments were filtered based on repeat regions, local entropy calculation and similarity between reference and alternative alignments. Larger than 3 unique alignments (at least one of them double stranded) were required to report a DNA fusion.

### Tumor Fraction

ctDNA fraction was estimated based on the mutant allele fraction of autosomal somatic mutations as described previously (Newman et al. 2016). Briefly, under the conservative assumption that each SNV may have loss of heterozygosity, the mutant allele fraction (MAF) and ctDNA fraction are related as MAF = (ctDNA * 1) / [(1 - ctDNA) * 2 + ctDNA *1], and so ctDNA = 2 / ((1 / MAF) + 1). Somatic mutations in genes with a detectable copy number change were omitted from ctDNA fraction estimation, thus only a subset of samples could have ctDNA fraction accurately estimated from mutation data.

### bTMB score estimation

Blood-based tumor mutational burden (bTMB) was defined as the number of somatic coding SNVs, including synonymous and nonsynonymous variants, within panel target regions. Because TMB estimation considers all variants (including synonymous and non-whitelist variants), higher variant call specificity is required. More stringent cut-offs were used for variant calls, and only variants with allele frequency ≥0.35% were used in score calculation. The bTMB score was weighted and normalized by the total effective targeted panel size within the coding region. 43 samples with the maximum somatic allelic frequency (MSAF) < 0.7% were excluded for bTMB estimation.

## REFERENCES

1. Soria F, Shariat SF, Lerner SP, et al: Epidemiology, diagnosis, preoperative evaluation and prognostic assessment of upper-tract urothelial carcinoma (UTUC). World J Urol 35:379–387, 2017

2. Ruvolo CC, Nocera L, Stolzenbach LF, et al: Incidence and survival rates of contemporary patients with invasive upper tract urothelial carcinoma. European urology oncology 4:792–801, 2021

3. Coleman JA, Yip W, Wong NC, et al: Multicenter Phase II Clinical Trial of Gemcitabine and Cisplatin as Neoadjuvant Chemotherapy for Patients With High-Grade Upper Tract Urothelial Carcinoma. Journal of Clinical Oncology 0:JCO.22.00763

4. Margulis V, Puligandla M, Trabulsi EJ, et al: Phase II Trial of Neoadjuvant Systemic Chemotherapy Followed by Extirpative Surgery in Patients with High Grade Upper Tract Urothelial Carcinoma. Journal of Urology 203:690–698, 2020

5. Subiela JD, Territo A, Mercade A, et al: Diagnostic accuracy of ureteroscopic biopsy in predicting stage and grade at final pathology in upper tract urothelial carcinoma: Systematic review and meta-analysis. Eur J Surg Oncol 46:1989-1997, 2020

6. Favaretto RL, Shariat SF, Savage C, et al: Combining imaging and ureteroscopy variables in a preoperative multivariable model for prediction of muscle-invasive and non-organ confined disease in patients with upper tract urothelial carcinoma. BJU Int 109:77–82, 2012

7. Margulis V, Youssef RF, Karakiewicz PI, et al: Preoperative multivariable prognostic model for prediction of nonorgan confined urothelial carcinoma of the upper urinary tract. J Urol 184:453–8, 2010

8. Petros FG, Qiao W, Singla N, et al: Preoperative multiplex nomogram for prediction of high-risk nonorgan-confined upper-tract urothelial carcinoma. Urol Oncol 37:292 e1–292 e9, 2019

9. Yoshida T, Kobayashi T, Kawaura T, et al: Development and external validation of a preoperative nomogram for predicting pathological locally advanced disease of clinically localized upper urinary tract carcinoma. Cancer Med 9:3733–3741, 2020

10. Baard J, de Bruin DM, Zondervan PJ, et al: Diagnostic dilemmas in patients with upper tract urothelial carcinoma. Nat Rev Urol 14:181–191, 2017

11. Kaag MG, O’Malley RL, O’Malley P, et al: Changes in renal function following nephroureterectomy may affect the use of perioperative chemotherapy. Eur Urol 58:581–7, 2010

12. Corcoran RB, Chabner BA: Application of Cell-free DNA Analysis to Cancer Treatment. N Engl J Med 379:1754–1765, 2018

13. Rose KM HH, Meeks J, et al.: Circulating and urinary tumour DNA in urothelial carcinoma—upper tract, lower tract, and metastatic disease. Nature Reviews Urology Ahead of print., 2023

14. Agarwal N, Pal SK, Hahn AW, et al: Characterization of metastatic urothelial carcinoma via comprehensive genomic profiling of circulating tumor DNA. Cancer 124:2115–2124, 2018

15. Barata PC, Koshkin VS, Funchain P, et al: Next-generation sequencing (NGS) of cell-free circulating tumor DNA and tumor tissue in patients with advanced urothelial cancer: a pilot assessment of concordance. Ann Oncol 28:2458–2463, 2017

16. Chalfin HJ, Glavaris SA, Gorin MA, et al: Circulating Tumor Cell and Circulating Tumor DNA Assays Reveal Complementary Information for Patients with Metastatic Urothelial Cancer. Eur Urol Oncol 4:310–314, 2021

17. Green EA, Li R, Albiges L, et al: Clinical Utility of Cell-free and Circulating Tumor DNA in Kidney and Bladder Cancer: A Critical Review of Current Literature. Eur Urol Oncol, 2021

18. Vandekerkhove G, Lavoie JM, Annala M, et al: Plasma ctDNA is a tumor tissue surrogate and enables clinical-genomic stratification of metastatic bladder cancer. Nature Communications 12, 2021

19. Christensen E, Birkenkamp-Demtröder K, Sethi H, et al: Early Detection of Metastatic Relapse and Monitoring of Therapeutic Efficacy by Ultra-Deep Sequencing of Plasma Cell-Free DNA in Patients With Urothelial Bladder Carcinoma. J Clin Oncol 37:1547–1557, 2019

20. Shohdy KS, Villamar DM, Cao Y, et al: Serial ctDNA analysis predicts clinical progression in patients with advanced urothelial carcinoma. Br J Cancer 126:430–439, 2022

21. Birkenkamp-Demtröder K, Nordentoft I, Christensen E, et al: Genomic Alterations in Liquid Biopsies from Patients with Bladder Cancer. Eur Urol 70:75–82, 2016

22. Flaig TW, Spiess PE, Agarwal N, et al: Bladder Cancer, Version 3.2020, NCCN Clinical Practice Guidelines in Oncology. J Natl Compr Canc Netw 18:329-354, 2020

23. Roupret M, Babjuk M, Burger M, et al: European Association of Urology Guidelines on Upper Urinary Tract Urothelial Carcinoma: 2022 Update. Eur Urol 79:62–79, 2021

24. Edge S: AJCC cancer staging manual. Springer 7:97-100, 2010

25. Zhang R, Zang J, Xie F, et al: Urinary Molecular Pathology for Patients with Newly Diagnosed Urothelial Bladder Cancer. J Urol 206:873–884, 2021

26. Fettke H, Kwan EM, Docanto MM, et al: Combined Cell-free DNA and RNA Profiling of the Androgen Receptor: Clinical Utility of a Novel Multianalyte Liquid Biopsy Assay for Metastatic Prostate Cancer. European Urology 78:173–180, 2020

27. Landrum MJ, Lee JM, Riley GR, et al: ClinVar: public archive of relationships among sequence variation and human phenotype. Nucleic Acids Res 42:D980–5, 2014

28. Adalsteinsson VA, Ha G, Freeman SS, et al: Scalable whole-exome sequencing of cell-free DNA reveals high concordance with metastatic tumors. Nat Commun 8:1324, 2017

29. Robin X, Turck N, Hainard A, et al: pROC: an open-source package for R and S+ to analyze and compare ROC curves. BMC Bioinformatics 12:77, 2011

30. Fujii Y, Sato Y, Suzuki H, et al: Molecular classification and diagnostics of upper urinary tract urothelial carcinoma. Cancer Cell 39:793–809.e8, 2021

31. Guo G, Sun X, Chen C, et al: Whole-genome and whole-exome sequencing of bladder cancer identifies frequent alterations in genes involved in sister chromatid cohesion and segregation. Nature Genetics 45:1459–1463, 2013

32. Audenet F, Isharwal S, Cha EK, et al: Clonal Relatedness and Mutational Differences between Upper Tract and Bladder Urothelial Carcinoma. Clinical Cancer Research 25:967–976, 2019

33. Lee JH, Saw RP, Thompson JF, et al: Pre-operative ctDNA predicts survival in high-risk stage III cutaneous melanoma patients. Annals of Oncology 30:815–822, 2019

34. Christensen E, Nordentoft I, Birkenkamp-Demtroder K, et al: Cell-free urine- and plasma DNA mutational analysis predicts neoadjuvant chemotherapy response and outcome in patients with muscle invasive bladder cancer. Clin Cancer Res, 2023

35. Robinson BD, Vlachostergios PJ, Bhinder B, et al: Upper tract urothelial carcinoma has a luminal-papillary T-cell depleted contexture and activated FGFR3 signaling. Nat Commun 10:2977, 2019

36. Sharma V, Miest TS, Juvet TS, et al: The Impact of Upper Tract Urothelial Carcinoma Diagnostic Modality on Intravesical Recurrence after Radical Nephroureterectomy: A Single Institution Series and Updated Meta-Analysis. J Urol 206:558–567, 2021

